# DNA methylation clock DNAmFitAge shows regular exercise is associated with slower aging and systemic adaptation

**DOI:** 10.1101/2022.07.22.22277842

**Authors:** Matyas Jokai, Ferenc Torma, Kristen M. McGreevy, Erika Koltai, Zoltan Bori, Gergely Babszki, Peter Bakonyi, Zoltan Gombos, Bernadett Gyorgy, Dora Aczel, Laszlo Toth, Peter Osvath, Marcell Fridvalszky, Timea Teglas, Balazs Ligeti, Regina Kalcsevszki, Aniko Posa, Sylwester Kujach, Robert Olek, Takuji Kawamura, Yasuhiro Seki, Katsuhiko Suzuki, Kumpei Tanisawa, Sataro Goto, Istvan Boldogh, Xueqing Ba, Dora Szabo, Kelvin J. A. Davies, Steve Horvath, Zsolt Radak

**Affiliations:** Research Institute of Sport Science, Hungarian University of Sport Science, Budapest, Hungary; Department of Biostatistics, Fielding School of Public Health, University of California Los Angeles, Los Angeles, CA 90095, USA; Faculty of Information Technology and Bionics, Pázmány Péter Catholic University, 1123 Budapest, Hungary; Interdisciplinary Excellence Center, Department of Physiology, Anatomy and Neuroscience, Faculty of Science and Informatics, University of Szeged, 6700 Szeged, Hungary; Department of Physiology, Faculty of Physical Education, Gdansk University of Physical Education and Sport, Gdańsk, Poland; Department of Athletics, Strength and Conditioning, Poznań University of Physical Education, Poznań, Poland; Faculty of Sport Sciences, Waseda University, Tokorozawa 2-579-15, Japan; Department of Microbiology and Immunology, University of Texas Medical Branch at Galveston, Galveston, TX 77555, USA; Key Laboratory of Molecular Epigenetics of Ministry of Education, Northeast Normal University, Changchun, Jilin, China; Institute of Medical Microbiology, Semmelweis University, Budapest, Hunagary; Ethel Percy Andrus Gerontology Centre of the Leonard Davis School of Gerontology; Division of Molecular & Computational Biology, Department of Biological Sciences, of the Dornsife College of Letters, Arts, and Sciences; and Department of Biochemistry & Molecular Medicine of the USC Keck School of Medicine, University of Southern California, Los Angeles, CA, 90089-0191, USA; Department of Human Genetics, David Geffen School of Medicine, University of California Los Angeles, Los Angeles, CA 90095, USA

## Abstract

DNAmPhenoAge, DNAmGrimAge, and the newly developed DNAmFitAge are DNA methylation (DNAm) based biomarkers that reflect the individual aging process. Furthermore, physical fitness is known to relate to the aging process, but its relationship to the gut microbiome has not yet been studied. Here, we examine the relationship among physical fitness, DNAm based biomarkers, and the microbiome in adults aged 33-88 with a wide range of physical fitness (including athletes with long-term training history). Higher levels of VO2max (ρ=0.2, p=6.4E-4, r=0.19, p=1.2E-3), Jumpmax p=0.11, p=5.5E-2, r=0.13 p= 2.8E-2), Gripmax (=ρ=0.17, p=3.5E-3, r=0.16, p=5.6E-3), and HDL levels (ρ=0.18, p=1.95E-3, r=0.19, p=1.1E-3) are associated with better verbal short term memory. In addition, verbal short term memory is associated with decelerated aging assessed with the new DNAm biomarker FitAgeAcceleration (ρ: −0.18, p=0.0017). DNAmFitAge is able to distinguish high fitness individuals from low/medium fitness individuals better than existing DNAm biomarkers and estimates a younger biological age in the high fit males and females (1.5 and 2.0 years younger, respectively). The microbiomal pathways are associated with VO2max, redox balance, and DNAmPhenoAge. PhenoAge Acceleration is influenced by pyruvate producing microbiomal pathways, where higher activity of these pathways lead to suppressed PhenoAge acceleration. Our research shows that regular physical exercise contributes to observable physiological, methylation, and microbiota differences which are beneficial to the aging process. DNAmFitAge emerged as a biological marker of the quality of life.

## Introduction

Aging is a natural process which depends on genetics, environment, and life-style factors. These factors make aging individualistic, and it has gained popularity in personalized medicine-especially through the use of epigenetic biomarkers. These biomarkers use a person’s DNA methylation to provide estimates of chronological age, biological age, rate of aging, mortality risk, etc. For example, DNAmPhenoAge and DNAmGrimAge are strong predictors of all-cause mortality and are associated with age-related diseases (Horvath and Raj, 2018; Lu et al., 2019). Furthermore, DNA methylation biomarkers have been able to capture environmental effects to aging. Monozygotic twins with different aging diseases have a different DNAmAge Acceleration (Fraga et al., 2005; Zhang et al., 2016), which underlines the significant role environmental and lifestyle factors have to aging phenotypes.

One of the striking effects of aging is a decrease in many physiological functions, and it has been shown that age-associated decline can be attenuated with regular physical activity (Booth et al., 2017). Regular exercise also decreases mortality risk and the incidence of age-related diseases including dementia, Alzheimer’s disease, osteoporosis, hypertension, cardiovascular diseases, cancer, stroke and arthritis (Radak et al., 2010). Moreover, regular physical activity has systemic effects on the body, influencing almost all of the organ function, cellular and organ metabolism, redox-sensitive cellular signaling, and activation of the immune system probably partly through the modulation of the microbiome (Radak et al., 2008a; Radak et al., 2008b; Quan et al., 2020).

This systemic adaptation provides reason for DNA methylation to be influenced by physical exercise and for physical activity to be a valuable component to DNAm-based aging biomarkers. However, only recently has physical activity been incorporated into a DNAm-based aging biomarker. DNAmFitAge, a new DNAm biomarker, provides an estimate of biological age using DNAm-based estimates of three physical fitness measurements: maximal oxygen uptake (DNAmVO2max), maximal gripping force (DNAmGripmax), and gait speed (DNAmGaitspeed) (McGreevy et al., 2022). While DNAmFitAge was validated in healthy adult populations, it was unknown how this new biomarker would perform in train in physical fit individuals or if it would capture epigenetic differences related to physical fitness.

Aging and physical activity are known factors that alter the bacterial flora in the gut microbiome (Reza et al., 2019). The microbiota of the gut is crucial for breaking down dietary nutrients, regulating intestinal and systemic immune responses, producing small molecules critical for intestinal metabolism, and generating several gasses that can modulate cellular function. Due to the complex function of the gut microbiome, the diversity of microbes is defined by the abundance of distinct organism types (Rook et al., 2017). However, the possible role or connection of the microbiome to epigenetic aging and physical fitness is still unknown. Therefore, we investigated the complex relationship among DNAm based biomarkers of aging, including DNAmFitAge, a variety of physiological functioning variables, blood serum measures including cholesterol, irisin level, and redox balance, and the microbiome on 303 healthy individuals aged between 33 and 88 years with a diverse level of physical fitness. Our research intends to show that regular physical exercise is related to microbiota and methylation differences which are both beneficial to aging and measurable.

## Methods

### Study population

The study was approved by the National Public Health Center in accordance with the Helsinki Declaration and the regulations applicable in Hungary (25167-6/2019/EÜIG). The subjects of this study were volunteers who signed a written consent form to participate in the investigation. A great number of volunteers (n=205) participated in the World Rowing Masters Regatta in Velence, Hungary, and 303 subjects total between 33-88 years old were included in the study. Subjects completed a questionnaire regarding their health, educational status, and life-style-including exercise habits. The master rowing group was very heterogeneous; many athletes had just one or two training sessions a week, while others had daily training. Therefore, we classified subjects into different fitness groupings based on the level of VO2max, which represents cardiovascular fitness. VO2max has been regarded as one of the best indicators of an athlete’s physical capacity (Hawley et al., 2014). Hence, subjects were divided into one of two fitness categories: medium-low fit group (MED-LOW FIT) (male n=50, female n=62) or highly fit (HIGH-FIT) group (male n=93 and female n=91) based on the 75th percentile of the VO2max values (Kaminsky et al., 2015) (Table 1.).

**Table 1.**
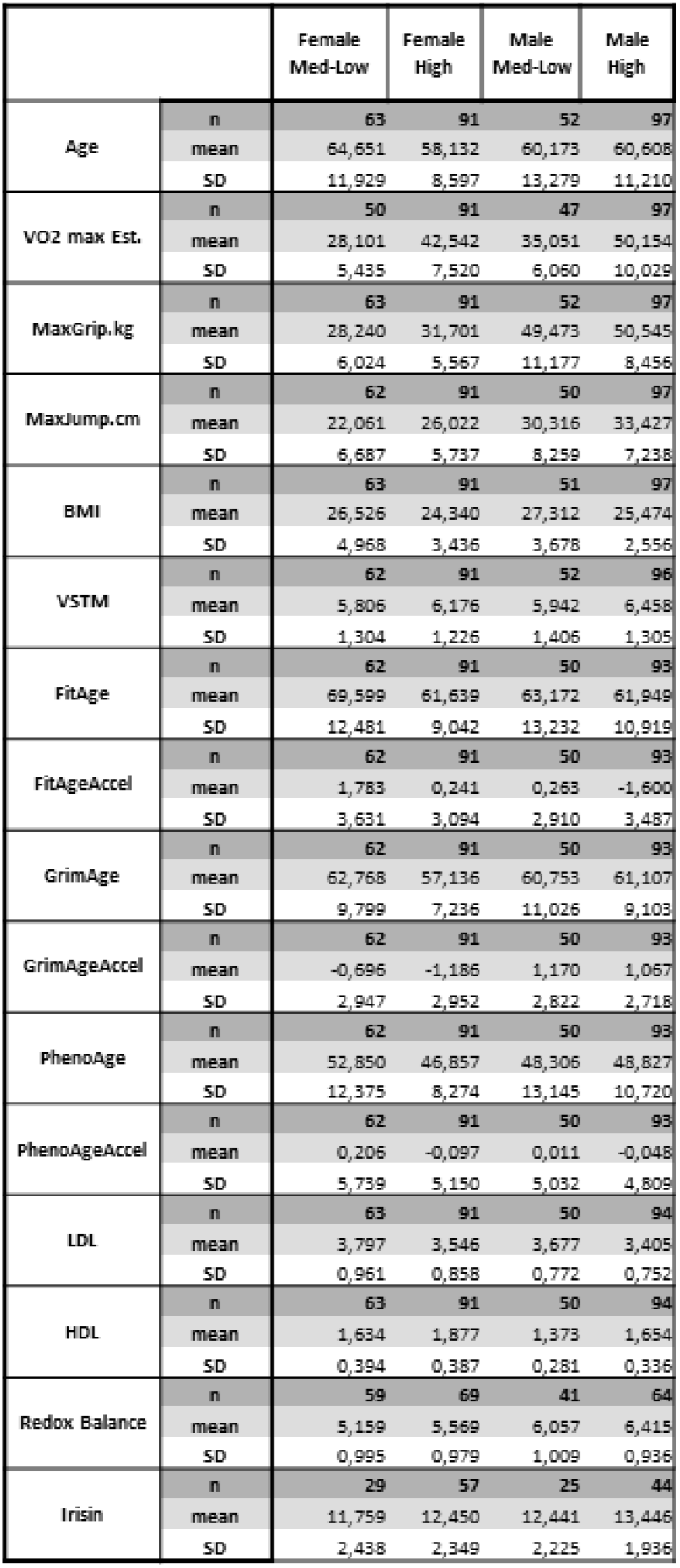
The characteristics and measured parameters of the subjects.

### Physiology tests

A detailed description of the methods used in this study can be found in the supplement. In brief, the digit span test was applied to assess the working memory (Martinez-Diaz et al., 2020), where larger values indicate better memory. Maximum hand gripping force is often used to measure age-associated decline in general muscle strength (Eika et al., 2019). The dynamic strength of the legs was assessed by the maximum vertical jump, using linear encoder (Lee et al., 1996). Body mass index was apprised by body composition monitor BF214 (Omron, Japan). Maximal oxygen uptake, VO2max, measures the volume of oxygen the body processes during incremental exercise in milliliters used in one minute of exercise per kilogram of body weight (mL/kg/min) and was measured through Chester step test (Izquierdo et al., 2019).

### Determination of hematologic and biochemical variables

Blood samples were collected before the subjects performed the VO2max evaluation test, and were stored in evacuated tubes containing EDTA as an anticoagulant for determination of erythrogram. Blood samples were centrifuged and stored at –80 C degrees. The erythrogram and biochemical tests were carried out in the Clinical Analysis Laboratory of Semmelweis University, Budapest.

### Measurement of Irisin

Plasma irisin was quantified using commercially available ELISA kits (EK-067–29, Irisin Recombinant, Phoenix Pharmaceuticals, Inc, Burlingame, USA). All samples from a particular subject were analyzed using the same plate (intra-assay). Intra- and inter-assay coefficients of variation were 4.1% and 15.2%, respectively.

### Assessment of Redox balance

The total amount of organic hydroperoxides in blood was spectrophotometrically estimated using the d-Roms (derivatives of reactive oxygen metabolites) test as described previously (Tsuchiya et al., 2008). In brief, 20 μL of blood was dissolved in an acetate-buffered solution (pH 4.8). The hydroperoxide groups react with the transition metal ions liberated from the proteins in the acidic medium and are converted to alkoxy and peroxy radicals. These newly formed radicals, the quantities of which are directly proportional to those of the peroxides present in serum, are trapped chemically with chromogen (N, N-dietyl-para-phenylendiamine), leading to formation of the corresponding radical cation. The concentration of this persistent species can be determined through spectrophotometric procedures (to detect its absorption at 505 nm). The concentrations are expressed in conventional units (Carratelli units; UCarr) in which 1 UCarr corresponds to 0.8 mg/L H_2_O_2_. The d-Roms test is performed using a FREE Carpe Diem analyzer (Wismerll CO, Ltd., Tokyo, Japan) (Tsuchiya et al., 2008).

### Measurement of Plasma Ferric-Reducing Ability

The plasma ferric-reducing ability was estimated using the biological antioxidant power test (BAP). In brief, ferric chloride is mixed with a special chromogen substrate, a thiocyanate derivative. Plasma (10 μL) prepared from each blood sample is added to this reaction mixture and incubated at 37°C for 5 min. The reduction of ferric ion is quantified by measuring the absorbance change at 505 nm. The BAP assays are also performed on a FREE Carpe Diem analyzer (Tsuchiya et al., 2008).

### Microbiome assay

Stool samples were collected for analysis of gut microbiota. Participants were instructed on proper methods for stool collection and all materials were provided in a convenient specimen collection kit. The samples were stored at −80 °C until further analysis. A frozen aliquot (200 mg) of each fecal sample was suspended in 250 ml of guanidine thiocyanate, 0.1 M Tris, pH 7.5, and 40 ml of 10% N-lauroyl sarcosine. Then, DNA extraction was conducted as previously described (Abraham et al., 2019). The DNA concentration and its molecular size were estimated by nanodrop (Thermo Scientific) and on agarose gel electrophoresis.

### Illumina sequencing

Fecal DNA has been used as input for the Illumina Nextera® XT DNA Sample Preparation Kit to construct indexed paired-end DNA libraries as previously described (Le Chatelier et al., 2013). DNA library preparation followed the manufacturer’s instruction (Illumina). We used the workflow indicated by the provider to perform cluster generation, template hybridization, isothermal amplification, linearization, blocking and denaturing and hybridization of the sequencing primers. The base-calling pipeline (version IlluminaPipeline-0.3) was used to process the raw fluorescent images and call sequences. We constructed one library (clone insert size 200 base pairs (bp)) for each of the first batch of 15 samples; two libraries with different clone insert sizes (135 and 400 bp) for each of the second batch of 70 samples, and one library (350 bp) for each of the third batch of 207 samples.

### Bioinformatics analysis

The quality of raw and trimmed reads was assessed with FastQC and MultiQC (Ewels et al., 2016). The low quality sequences were filtered and trimmed by Trimmomatic (Bolger et al., 2014) and only sequences with minimal length of 36 and low quality base calls were discarded (phred score <30). The host contamination (reads aligning to human reference genome (bowtie2 v2.4.2 (Langmead and Salzberg, 2012), GRCh38)) were discarded. The taxonomic characterization was carried out using the MetaPhlAn3 (Beghini et al., 2021) and pathway abundance and other molecular function profiles (such as GO) were estimated with HUMAnN3 pipeline. The rare (present maximum 10% in all samples) and low abundance (support of less than 0.01% abundance) taxa were discarded from the subsequent analysis. After the filtering process, a Bayesian-Multiplicative replacement of zeros was carried out using the z Composition R package, which was followed by central log-ratio (CLR) transformation of count and ratio values as implemented in scikit-bio.

### Measurement of DNA Methylation

Epigenome wide DNA methylation 85K was measured with the Infinium MethylationEPIC BeadChip (Illumina Inc., San Diego, CA) according to the manufacturer’s protocol. In short, 500 ng of genomic DNA was bisulfite converted using the EZ-96 DNA Methylation MagPrep Kit (Zymo Research, Irvine, CA, USA) with the KingFisher Flex robot (Thermo Fisher Scientific, Breda, Netherlands). The samples were plated in randomized order. The bisulfite conversion was performed according to the manufacturer’s protocol with the following modifications: For binding of the DNA 15 µl MagBinding Beads was used. The conversion reagent incubation was done according to the following cycle protocol: 16 cycles of 95°C for 30 seconds followed by 50 °C for 1 hour. After the cycle protocol the DNA was incubated for ten minutes at 4 °C. Next, DNA samples were hybridized on the Infinium MethylationEPIC BeadChip (Illumina Inc., San Diego, CA) according to the manufacturers protocol with the modification that 8 µl bisulfite treated DNA was used as start material.

Quality Control of the DNA methylation data was performed using minfi, Meffil and ewastools packages with R version 4.0.0. Samples which failed technical controls, including extension, hybridization and bisulfite conversion, according to the criteria set by Illumina, were excluded. Samples with a call rate < 96% or at least with 4% of undetected probes were also excluded. Probes with a detection p-value > 0.01 in at least 10% of the samples were set as undetected. Probes with a bead number < 3 in at least 10% of the samples were excluded. We used the “noob” normalization method in R to quantify methylation level (Triche et al., 2013). The details on the processing of DNAm data and the calculation of the measures of aging, or pace of aging, were calculated using Horvath’s online age calculator (https://dnamage.genetics.ucla.edu/).

### Epigenetic biomarkers

The development of epigenetic clocks is reviewed in Horvath and Raj (Horvath and Raj, 2018). Epigenetic clocks are considered highly promising molecular biomarkers of aging (Jylhava et al., 2017). The most commonly used epigenetic clocks are Hannum’s blood-specific clock (Hannum et al., 2013), and Horvath’s pan-tissue clock (Horvath, 2013), which are based on levels of DNAm at 71 and 353 CpG sites, respectively. These clocks are highly correlated with chronological age, and the discrepancy resulting from the regression of DNAm age on calendar age—referred to as epigenetic age acceleration—is associated with increased risk of all-cause mortality (Marioni et al., 2015; Chen et al., 2016). The first-generation epigenetic clock such as the pan tissue clock from Horvath 2013 exhibit statistically significant but relatively weak associations with clinical biomarkers and mortality risk. Far stronger associations with mortality risk and a host of age related conditions can be observed with so-called second-generation epigenetic clocks such as PhenoAge (Levine et al., 2018) and GrimAge (Lu et al., 2019). So far GrimAge appeared to be the most predictive model for mortality risk estimator (McCrory et al., 2021)5).

DNAmFitAge represents a new epigenetic biomarker that incorporates physical fitness. The creation of DNAmFitAge was recently described (Lu et al., 2019). In short, DNAm fitness biomarkers were created using LASSO penalized regression (DNAmGaitspeed, DNAmGripmax, DNAmFEV1, and DNAmVO2max). The DNAm fitness biomarkers for gaitspeed, gripmax, and FEV1 are built for either males or females and have two versions each. One version uses chronological age and DNA methylation to form an estimate of the fitness parameter, and the other version uses DNA methylation only to form estimates. We evaluate both the age and no age versions. DNAmVO2max uses age as a covariate and is built for both sexes. DNAmFitAge combines four DNAm-based biomarkers variables: three of the DNAm fitness biomarkers: DNAmGripmax noAge, DNAmGaitSpeed noAge, and DNAmVO2max, and DNAmGrimAge, a biomarker of mortality risk (Lu et al., 2019). Finally, FitAgeAcceleration is the age-adjusted estimate of DNAmFitAge formed from taking the residuals after regressing DNAmFitAge onto chronological age. FitAgeAcceleration provides an estimate of epigenetic age acceleration, ie how much older or younger a person’s estimated biological age is from expected chronological age.

### Statistical Analysis

The relationship between target and predictor variables were evaluated using multiple linear regression controlling for age and sex. Analyses were conducted using Statistica 13 software (TIBCO). For analysis of irisin, a variable for plate was included to control for possible batch effects. Fitness group differences were investigated by two-way ANOVA using sex and fitness group as factors; group means were compared by Tukey’s HSD. If data did not follow the normal distribution, assessed by Shapiro-Wilks test, the Kruskal–Wallis test was applied instead. The association between verbal short-term memory and biochemical/physiological markers were analyzed by calculating Spearman’s rho and Kendall’s tau.

### Physical Fitness to DNAm Biomarkers

We used two sample t-tests and non-parametric Kruskal-Wallis tests to determine if DNAm biomarkers were significantly different between the high fitness and low-med fitness groups in males and females (Table 1). We use the age-adjusted DNAm variables (FitAge Acceleration, GrimAge Acceleration, and PhenoAge Acceleration) to remove any age effect seen between groups. The same t-tests and Kruskal-Wallis tests were performed for physical fitness parameters Gripmax and Jumpmax (relative and absolute) to provide a reference for the DNAm-based surrogates.

VO2max is excluded from the table because VO2max was used to form high fit and low-med fitness groups. Furthermore, DNAmVO2max is excluded from the table because subjects from the study were used to construct the DNAmVO2max biomarker so observed differences by group is simply an artifact of the training dataset.

### Microbiome Analysis

The compositional similarities between the different groups were investigated with PERMANOVA and the differential abundance testing was done using the Wilcoxon signed-rank test and the Wilcoxon rank-sum test.

## Results

### Age-related physiological functioning and blood markers

Aging resulted in a decline in all measured physiological functions in MED-LOW FIT and HIGH-FIT groups (Figure 1). The rate of decline may be slower for high-fit groups, especially at older ages, but only the age associated decline in Jumpmax differs by fitness level. Interestingly, Jumpmax, which is used to evaluate anaerobic power (Vandewalle et al., 1987), is the only measured physiological function in which the decline is attenuated by fitness level, where high fit individuals have a slower decline. The change in LDL, HDL, and Redox balance by physical fitness and sex are shown in Figure 2. HDL appears constant in males across age in either fitness group, and the high-fit males have consistently higher HDL levels than med-low fit males. The top panel of Figure 3 presents the coefficients and p-values from multiple linear regression for serum irisin levels; irisin levels decrease with age (0.23 average decrease for every 1 year older). Furthermore, Figure 3 shows HDL is positively associated with irisin and that HDL is significantly different between high fit and low/med fit males and females.

**Figure 1.**
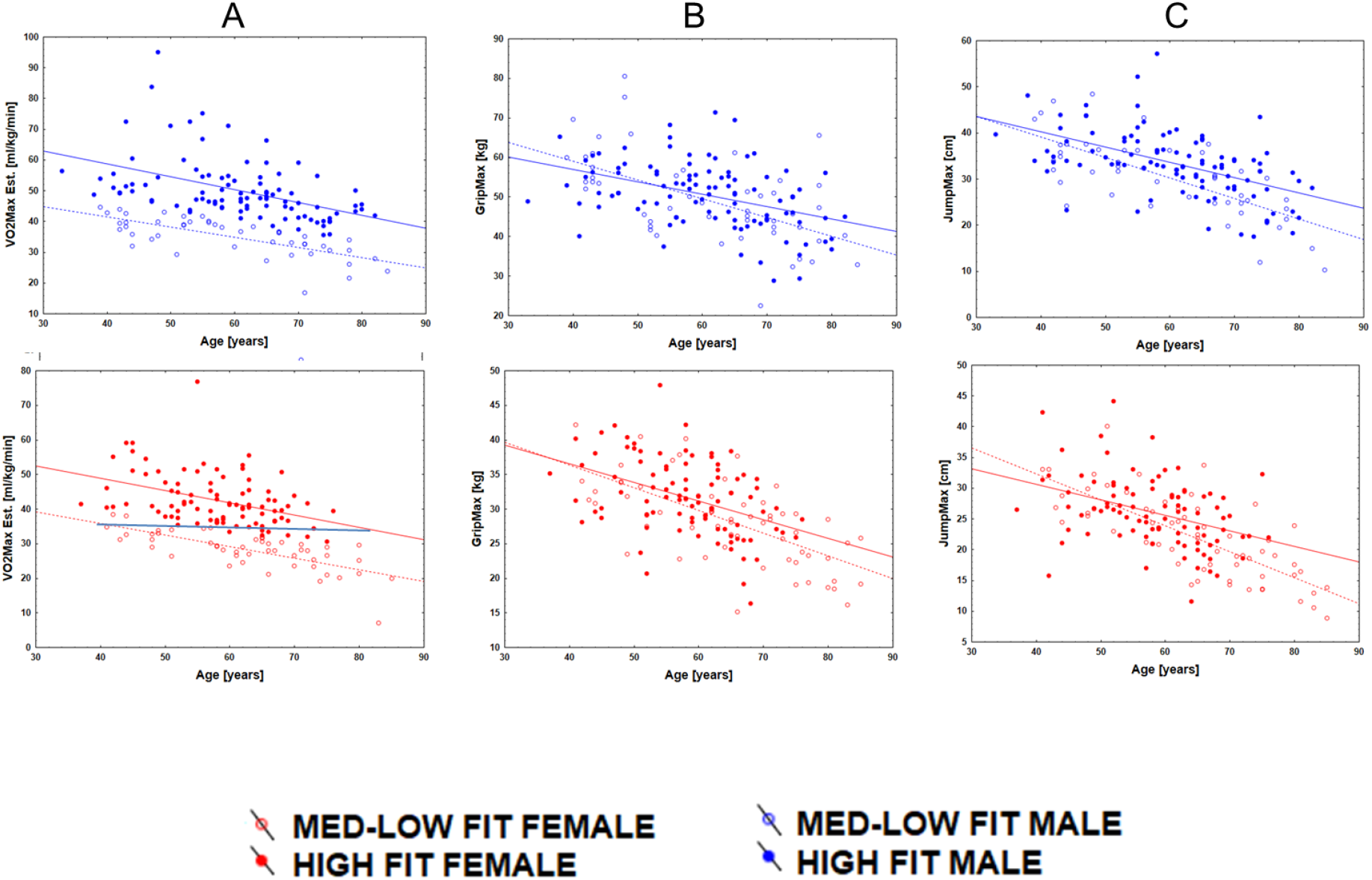
Age related decline in VO2max (A), GripMax (B) and JumpMax (C) of male and female HIGH-FIT and MED-LOW-FIT subjects. Regardless of fitness level VO2max, GripMax and JumpMax decreased as a result of aging, however the decrease started from higher values and subjects from HIGH-FIT group reached the values of MED-LOW-FIT subjects more than 20 years older age.

**Figure 2.**
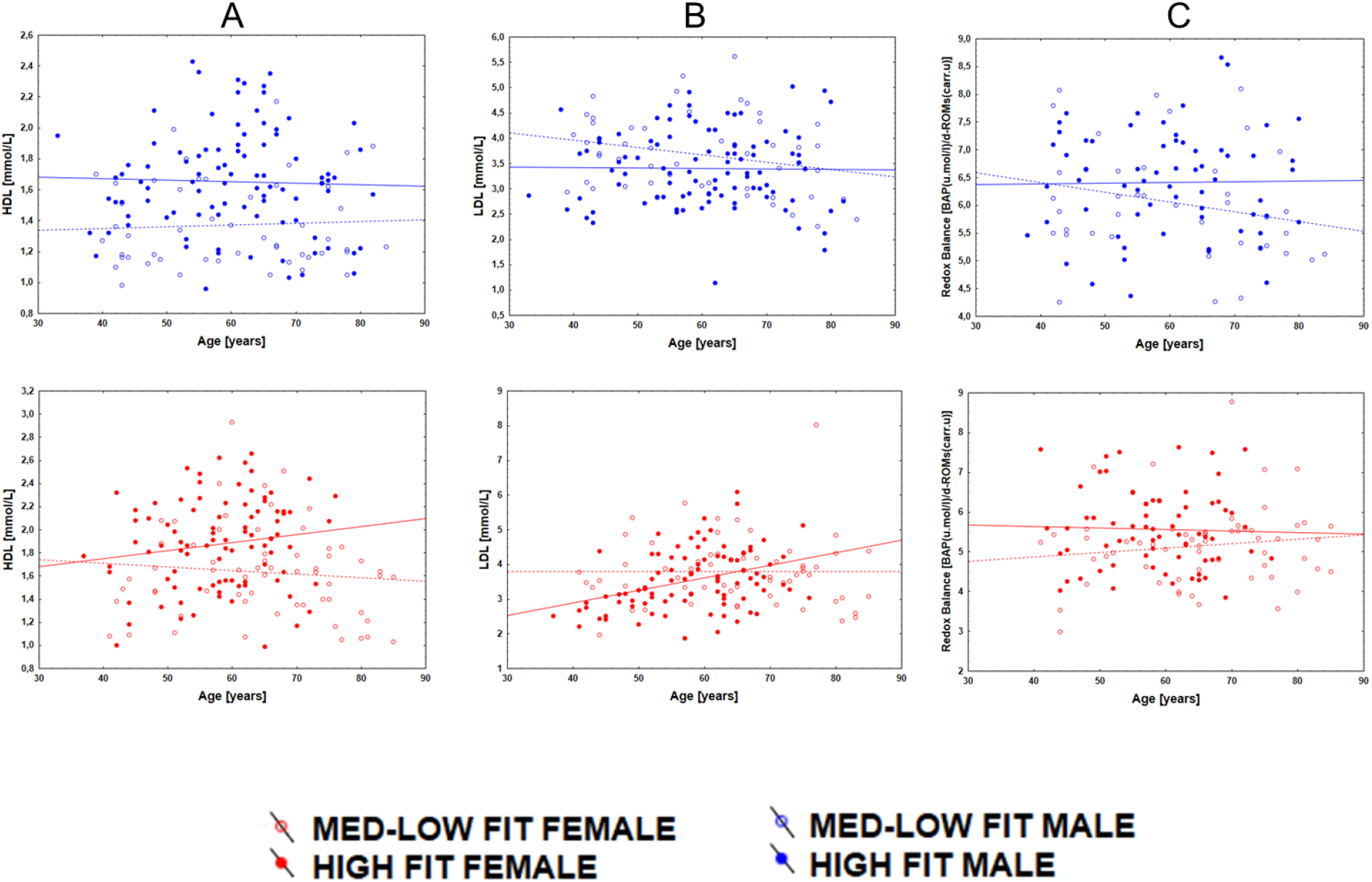
Age-associated change in HDL (A), LDL (B) and Redox Balance (C) of male and female HIGH-FIT and MED-LOW-FIT subjects. At HIGH-FIT males the HDL level was higher than in MED-LOW-FIT group and age association changes were not significant. On females, both HDL and LDL levels tend to increase with aging. The Redox balance remained unaltered by aging in both gender.

**Figure 3.**
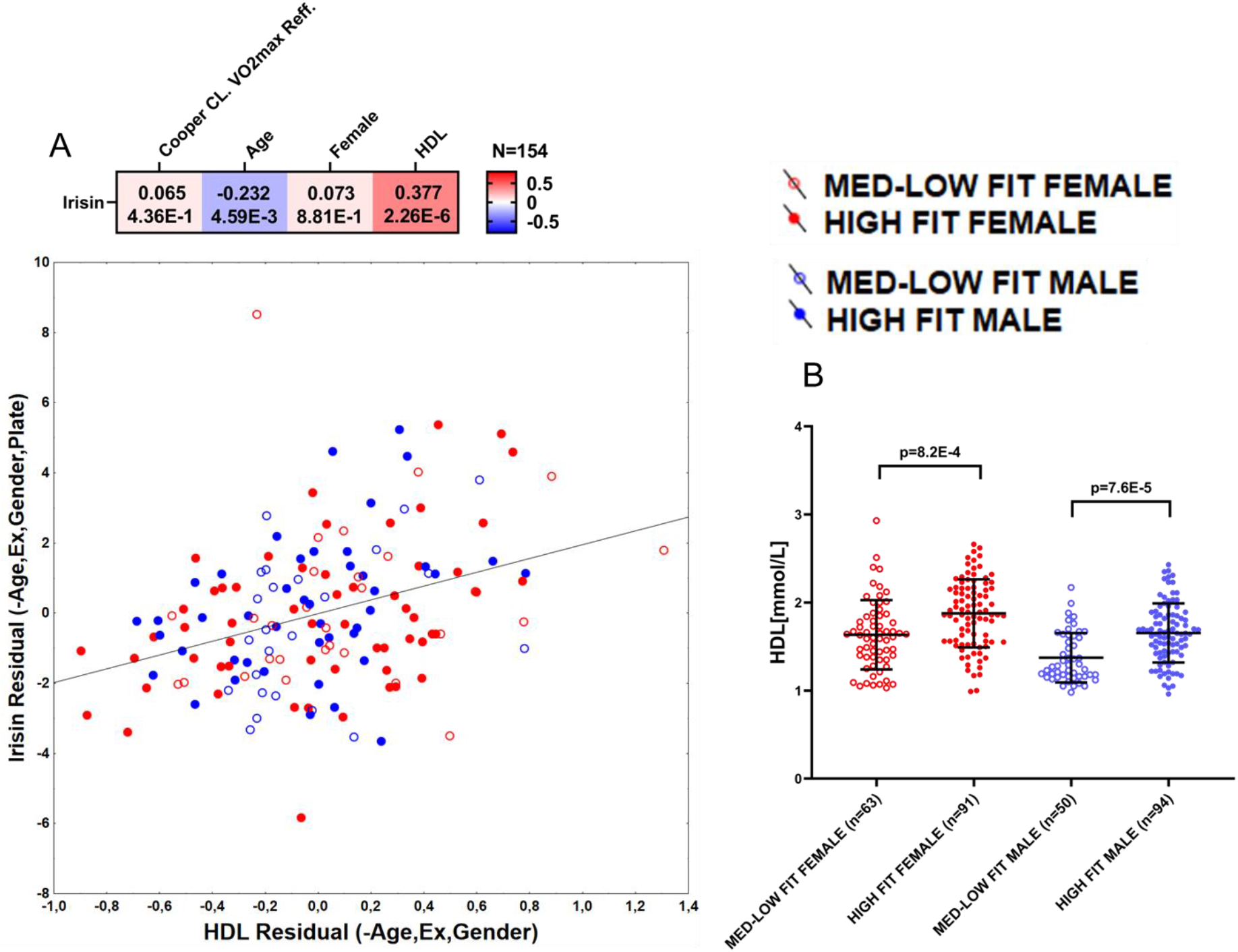
Correlation between HDL and irisin (A) and fitness and gender associated levels of HDL (B). Heath-map shows the correlation between irisin and HDL levels adjusted for age, gender, fitness level and batch (data not shown) with corresponding partial r (r_p_) and p values. Cartesian graph is showing the residuals of Irisin∼Age+Gender+Fitness level + plate model against residuals of HDL ∼Age+Gender+Fitness level model. Grouped plots are showing data distribution and group differences in HDL levels.

Better short term memory is observed in younger, leaner, and more physically fit individuals (Figure 4). The digit span test is the only physiological test independent of sex, which provides a measurement of verbal short term memory. Higher levels of VO2max (p=0.0013), JumpMax (p=0.028), GripMax (p=0.0056), as well as HDL levels (p=0.0011) were associated with better verbal short memory. Importantly, the newly created FitAgeAcceleration is also negatively correlated with digit span test results (p=0.0002), meaning better verbal memory is associated with decelerated aging.

**Figure 4.**
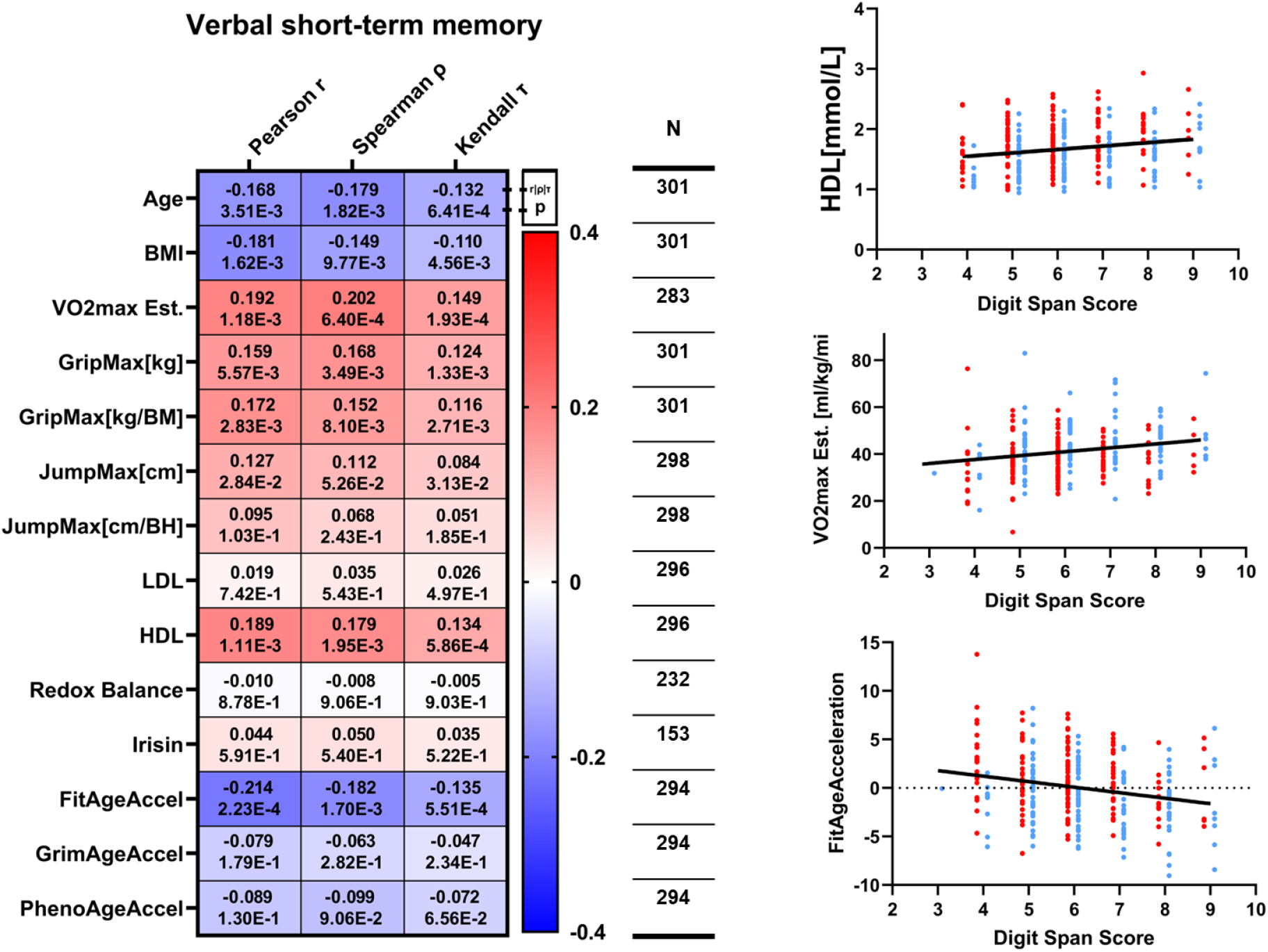
The association between verbal short term memory and physiological, biochemical and DNAm based parameters. On the heath-map Pearson’s r, Sperman’s rho and Kendal’s tau and corresponding p values are presented. The scatter-plots are sowing digit snap scores in respect of marked physiological parameters, red dots mark female, blue dots mark male subjects.

The relationship between physiological, biochemical, and DNAm biomarkers to age, sex, and physical fitness were as expected (Figure 5). All of the DNAm based biomarkers: DNAmFitAge, DNAmGrimAge, and DNAmPhenoAge have a strong relationship with chronological age (p < 1.0E-35). The measured physiological tests: VO2max, JumpMax, and GripMax have negative association with age and females have a lower mean value than males. Importantly, high fit individuals (classified through VO2max) are associated with younger DNAmFitAge (p=2.6E-5), lower BMI (p=9.7E-6), stronger relative GripMax (p=1.6E-4), farther JumpMax (p=2.6E-4), higher HDL levels (p=3.4E-9), and higher redox balance (p=4.3 E-3).

**Figure 5.**
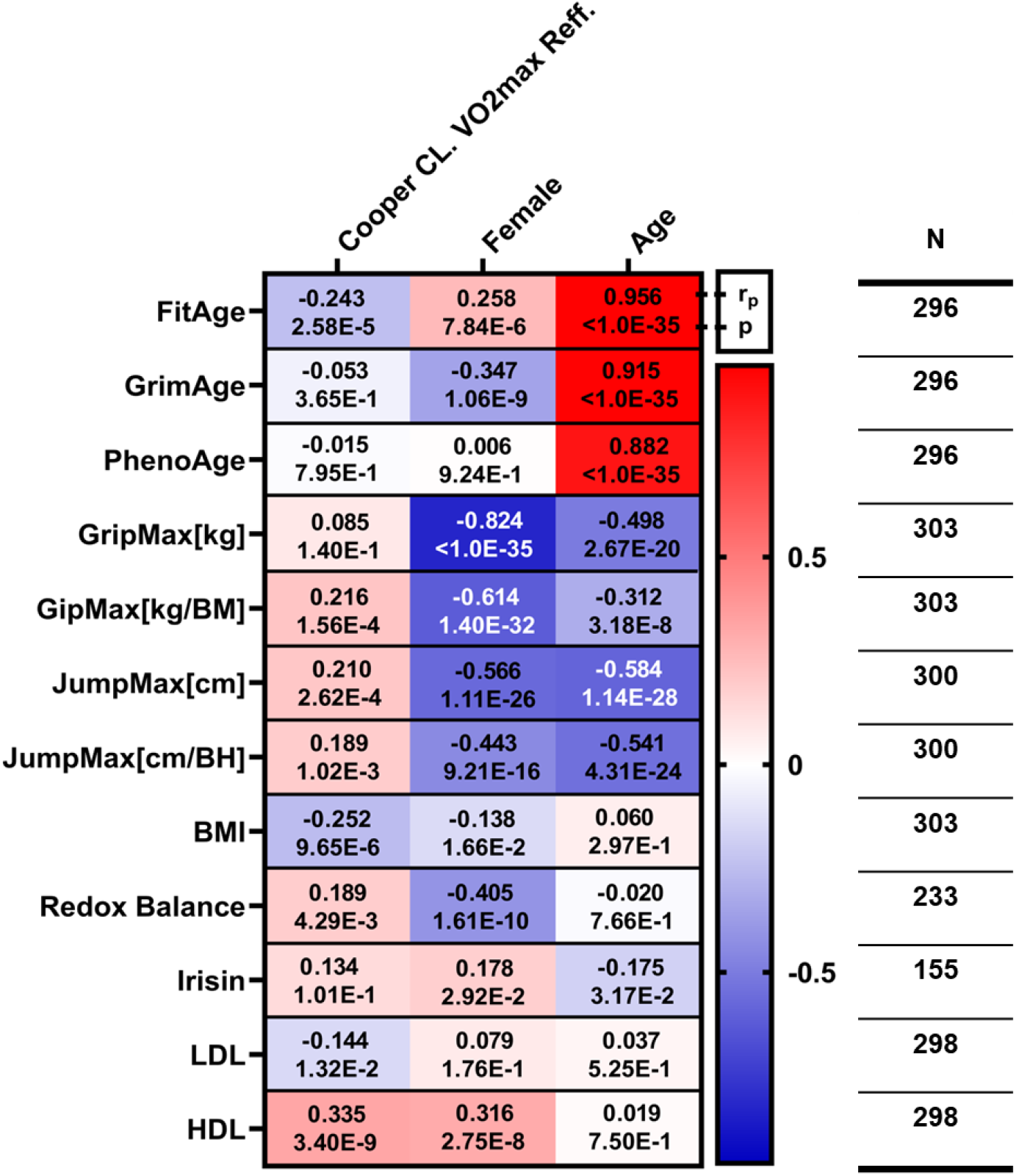
The relationship measured and calculated parameters with fitness level, gender and age. Results of the multiple linear regression models can be found in each cells. Row titles are the dependent variables, column titles are independent variables in the model. Each row represents a multiple lineal regression model and cells are containing partial correlation coefficients (r_p_) and corresponding p-values. N values at the end of each rows are showing the available sample size for all 4 variables in the model.

### DNAmFitAge and other DNAm biomarkers

FitAge Acceleration had the most powerful relationship to physical fitness parameters, BMI, and blood serum markers compared to GrimAge and PhenoAge Acceleration (Figure 6). Moreover, the direction of effect is as expected. A positive FitAge Acceleration corresponds to an older estimated biological age than your true chronological age (+ = older), whereas a negative FitAge Acceleration indicates being estimated to be younger than you are (- = younger / fitter). For every 1 year older FitAge Acceleration is, there is an average 0.29 decrease in relative grip strength (kg force/ body mass), 0.12 cm decrease in relative jumping distance (cm distance/ body height), 0.32 increase in body mass index, 0.31 decrease in blood HDL, 0.28 decrease in redox balance and 0.17 increase in blood irisin. The direction of effect is generally conserved across FitAge, GrimAge, and PhenoAge Acceleration, but the significance and magnitude of effect is stronger with FitAge Acceleration.

**Figure 6.**
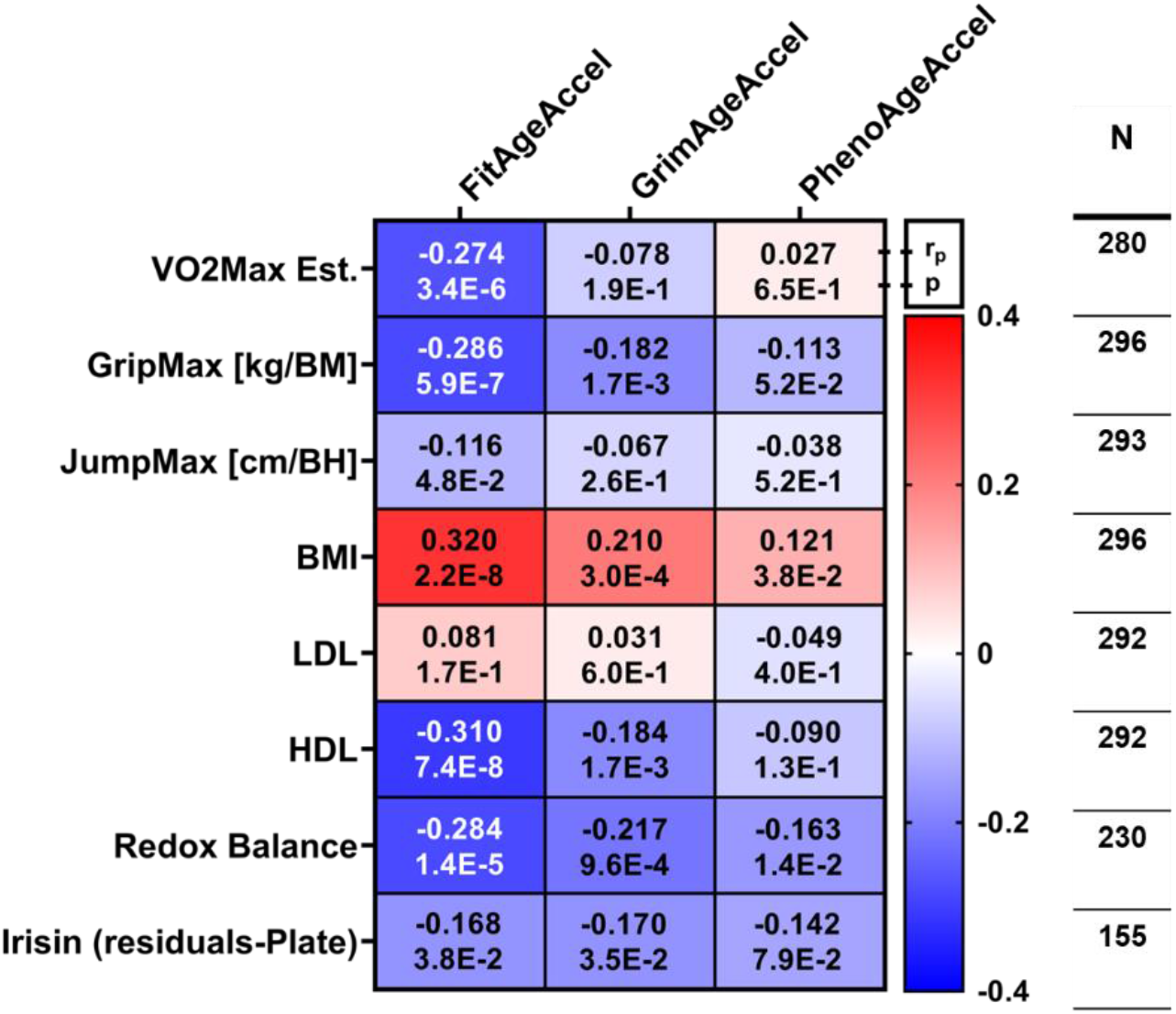
The relationship between physiological, biochemical parameters with acceleration of FitAge, GrimAge and PhenoAge. Results of the multiple linear regression models can be found in each cells adjusted for gender. Each row (dependent variable) and column (independent variable) intersection represents a multiple lineal regression model (with gender variable that is not shown) and cells are containing partial correlation coefficients (r_p_) and corresponding p-values. N values at the end of each rows are showing the available samples for all 3 in the model. For Irisin batch adjusted residual values were included for the multiple regression model.

FitAge Acceleration, but not GrimAge or PhenoAge Acceleration, distinguishes high fit subjects from low/med fit subjects in males and females (Table 1, Figure 7). FitAge Acceleration is 1.5 years younger on average in high fit females compared to low-med fit females (p=0.005), and FitAgeAcceleration is 2.0 years younger on average in high fit males compared to low-med fit males (p=0.0007). GrimAge Acceleration and PhenoAge Acceleration estimate younger values in the high-fit groups of males and females, but neither are significantly different (0.25 < p < 0.82). Furthermore, the differences observed between female fitness groups with GrimAge and PhenoAge Acceleration have smaller magnitudes (0.5 and 0.2 respectively) than FitAge Acceleration. Therefore, highly fit females and highly fit males are estimated to be 1.5 and 2.0 years biologically younger on average than their low to medium fit counterparts, suggesting regular physical exercise is protective to biological age in males and females.

**Figure 7.**
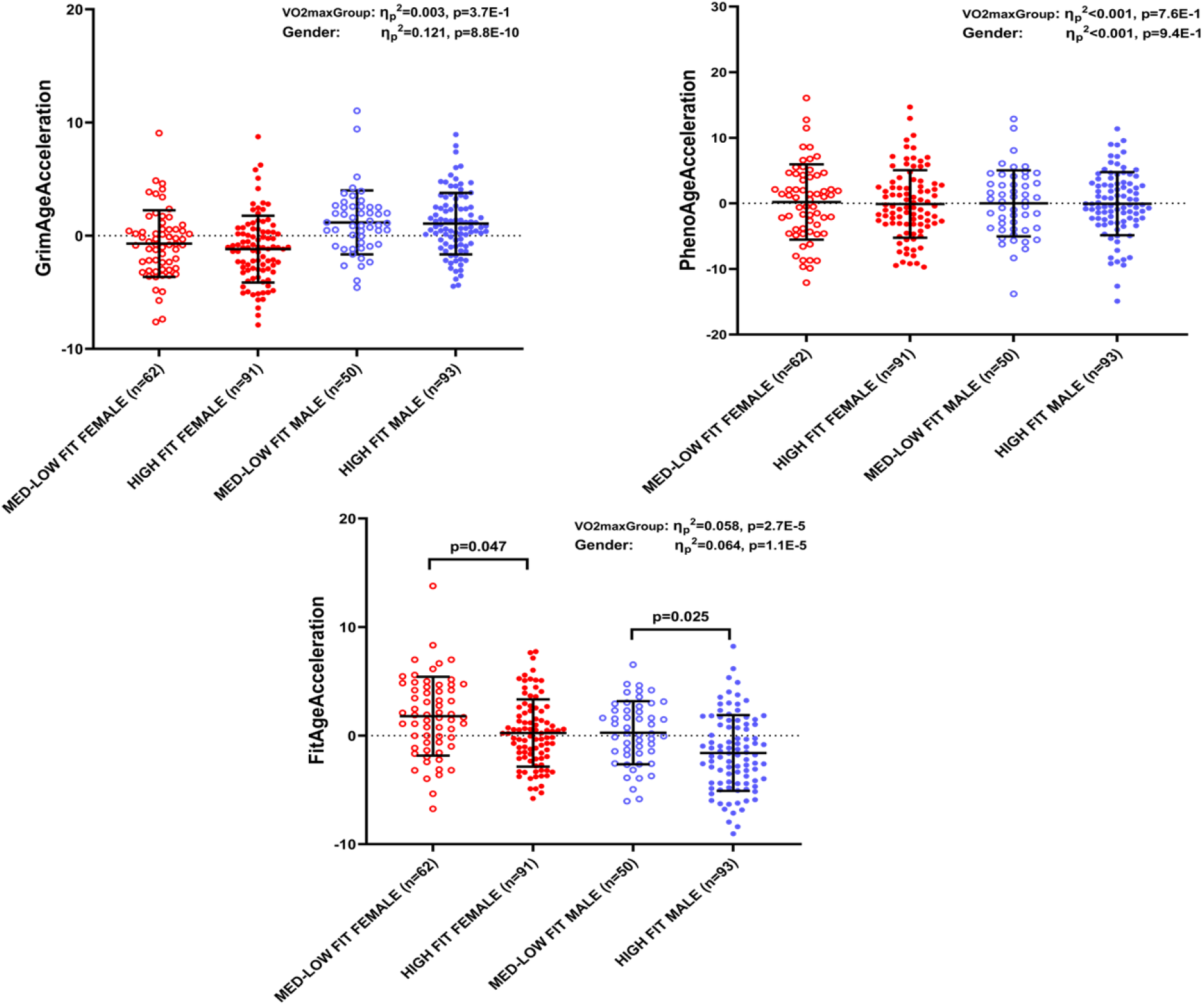
The Age Acceleration of DNA methylation based biological age estimators in the studied population. For effect size partial eta squared values (ηp2) are shown on the grouped plots with matching p values.

### DNAm Fitness Biomarkers

The underlying DNAm fitness biomarkers DNAmGripmax, DNAmGaitSpeed, and DNAmFEV1 can distinguish high-fit females from low-med fit females, but do not distinguish fitness grouping in males. Regardless if chronological age is used in the DNAm fitness biomarker construction, each biomarker has similar estimated differences in female fitness groups: 1.49 and 1.44 for DNAmGripmax, 0.10 and 0.09 for DNAmGaitspeed, and 0.14 and 0.15 for DNAmFEV1. We hypothesize the DNAm fitness biomarkers could not detect differences in the male fitness groups because the true physical fitness parameters are not very distinct between the male groups (relative gripmax p=0.049, jumpmax p=0.070).

### Microbiome Pathway

A number of fecal microbiota metabolic pathways are associated with VO2max, redox balance, PhenoAge and FitAge Acceleration (Figure 8). Positive association was found between VO2max and pathways that are related to carbohydrate and fat metabolism, L-arginine biosynthesis II acetyl cycle, L-arginine biosynthesis I via L-ornithine, flavin biosynthesis and glyoxylate cycle. Redox balance was positively associated to pyruvate fermentation to propanoate and negatively associated to menaquinol 11 biosynthesis super pathways. PhenoAge Acceleration negatively correlated to galacturonate degradation, D-fructuronate degradation, and D-glucuronosides degradation controlling microbiomal pathways. FitAge Acceleration was associated with L-isoleucine biosynthesis I from threonine.

**Figure 8.**
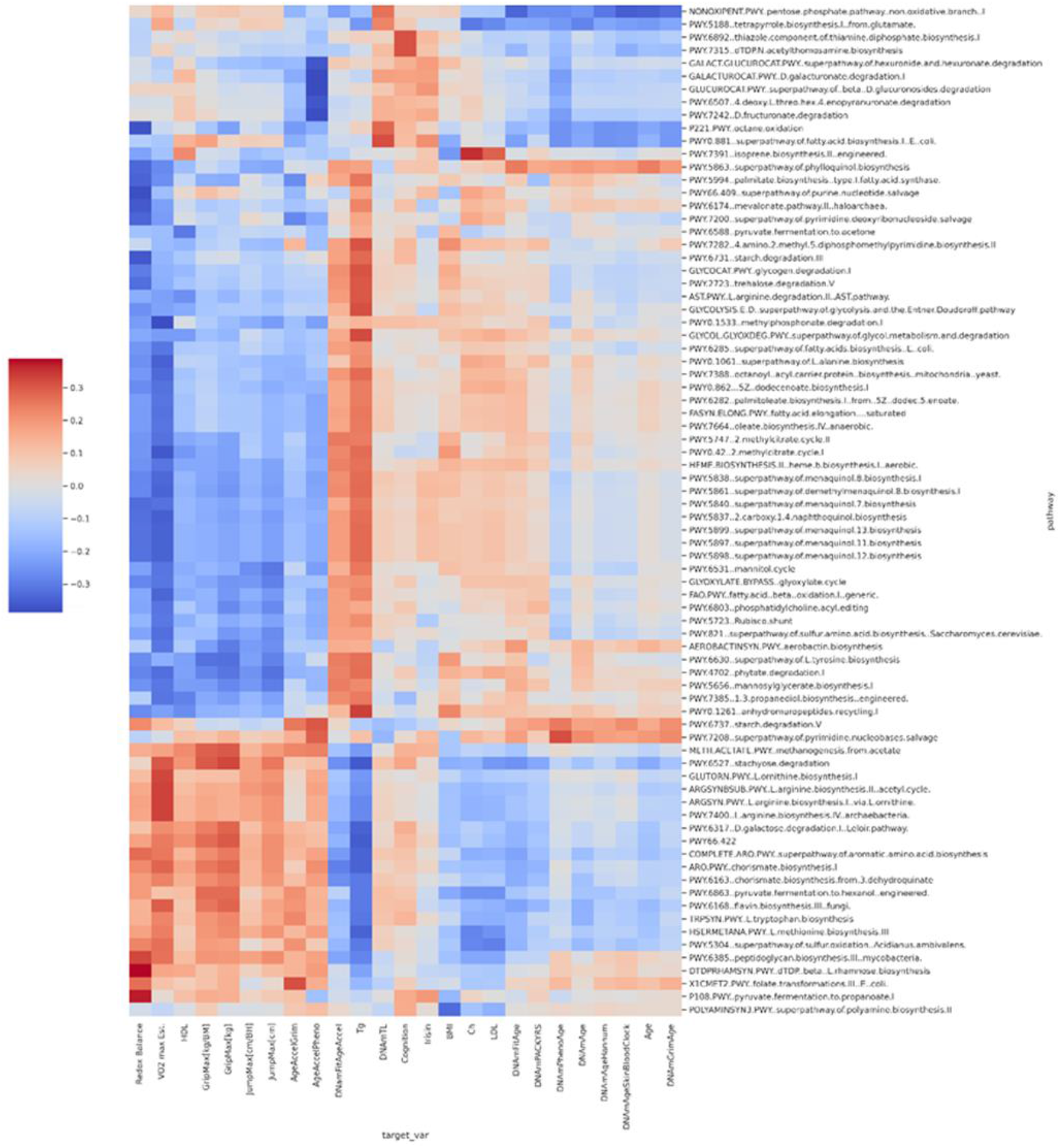
Heat map on the relationship between physiological function and epigenetical age markers and microbiome related molecular pathways. Positive association was found between VO2max and L-arginine biosynthesis II acetyl cycle, L-arginine biosynthesis I via L-ornithine, and flavin biosynthesis. Redox balance was positively associated to pyruvate fermentation to propanoate and negatively associated to menaquinol 11 biosynthesis super pathways. PhenoAge Acceleration negatively correlated to galacturonate degradation, D-fructuronate degradation, and D-glucuronosides degradation controlling microbiomal pathways. FitAge Acceleration was associated with L-isoleucine biosynthesis I from threonine.

Fecal microbiotal gene diversity via the Shannon diversity index may appear more complex in the high fit group compared to the med-low group, however the difference was not significant (species level p = 0.204, family level p=0.5, Figure. 9). Four microbial pathways were found to differ by physical fitness between high fit and med-low fit groups. The microbial pathways found to be significantly different in high fit and low-med groups were PWY-5484 Gylcolysis, PWY: 7234 Inosine 5’ biosynthesis, PYRIDNUCSAL-PWY: NAD salvage, and PWY 5676: Acetyl-CoA fermentation pathways (Figure 10).

**Figure 9.**
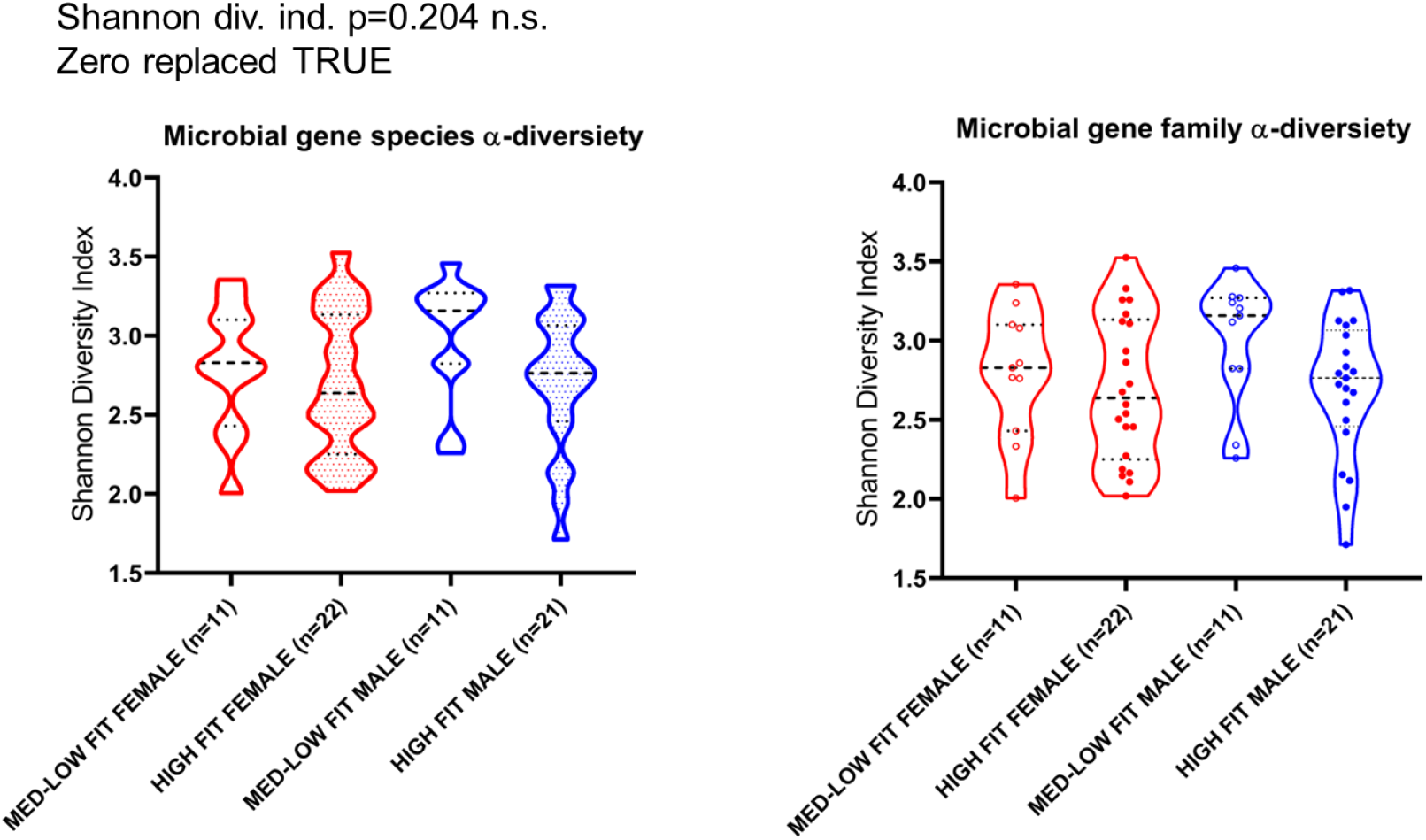
Shannon Diversity Index of gene in the species and family level. HIGH-FIT group tend to have higher diversity in species level p = 0.204, and in family level p=0.5, than MED-LOW-FIT group, but the difference did not reach the significant level. HIGH-FIT male N=21 HIGH-FIT female N=22, MED-LOW-FIT male N=11, MED-LOW-FIT female N=11.

**Figure 10.**
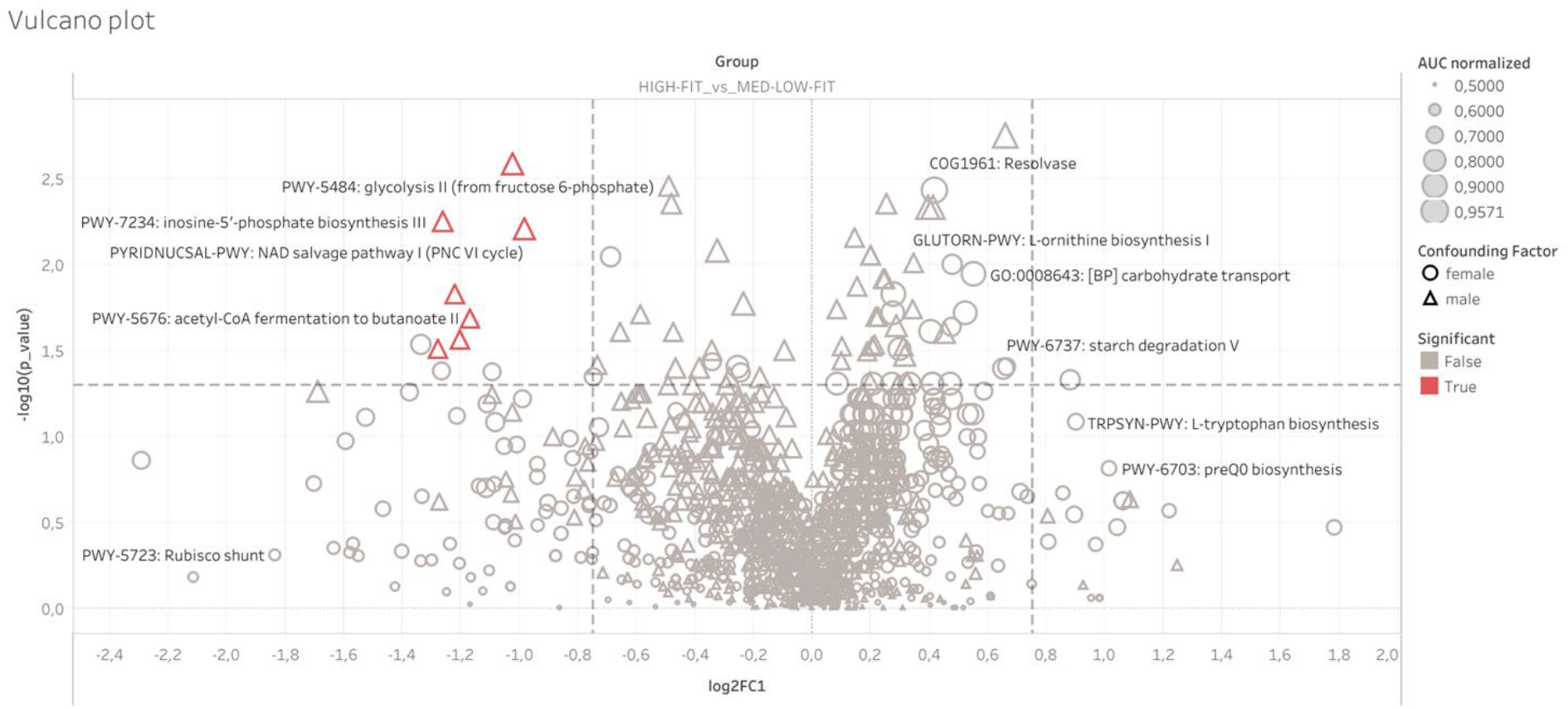
The relationships between microbiome related molecular pathways with the level of physical fitness. Fecal microbiota metabolic pathways are associated with VO2max, redox balance, and FitAge Acceleration (p<0.05). Positive association was found between VO2max and L-arginine biosynthesis II acetyl cycle, L-arginine biosynthesis I via L-ornithine, and flavin biosynthesis.

## Discussion

Lifestyle choices including healthy nutrition and regular exercise reduce mortality risk, and arguably slow the aging process. Here, we evaluated the new DNA methylation based biomarker, DNAmFitAge, which relates physical exercise to the epigenome, in healthy and athletic adults. DNAmFitAge along with previously published epigenetic mortality risk estimators DNAmGrimAge and DNAmPhenoAge predict coronary heart disease risk, comorbidities, and disease free status (McGreevy et al., 2022). However, we demonstrate FitAge Acceleration is better able to capture both general and individual methylation-based alterations from exercise-induced adaptations. FitAge Acceleration has a stronger relationship with verbal memory, HDL, BMI, Redox Balance, VO2max, JumpMax and GripMax.

The present investigation revealed that long-term regular exercise attenuates the age-associated decline in physiological function, including verbal short term memory, overall body strength, explosive strength, and cardiovascular fitness. This may suggest that individuals with higher levels of physiological function could have a higher physiological quality of life and a decreased risk of mortality (Kyu et al., 2016). Furthermore, DNAmFitAge shows higher levels of physiological functioning correspond to decelerated aging. FitAge Acceleration estimates high fit individuals have a 1.5 to 2.0 younger biological age on average compared to low-med fit individuals in females and males, respectively. These findings further support the hypothesis that regular exercise is protective to health and beneficial to the aging process.

HDL and irisin have complex roles in physiology, and the positive relationship we observe between physical exercise and HDL and irisin align with the protective effects seen between HDL and irisin with glucose homeostasis. HDL is involved in atheroprotection, inflammation, oxidative stress, nitric oxide production, and regulation of plasma glucose homeostasis (Constantinou et al., 2016). Infusing HDL into skeletal muscle can help control glucose uptake in people with type 2 diabetes mellitus (Drew et al., 2009). Additionally, C2C12 cells treated with irisin helped protect against high glucose cytotoxicity and preserved crucial AMPK-insulin receptor signaling (Yano et al., 2020). This molecular interaction may potentially explain the protective effects of exercise on higher HDL and irisin to glucose homeostasis.

Furthermore, higher HDL has a protective role in neurodegenerative diseases (Robert et al., 2021), and exercise has been shown to have a neuroprotective effect (Radak et al., 2010; Quan et al., 2020). In this study, we describe a protective association between HDL levels, VO2max, and a digit memory test. In addition, we find that younger FitAge Acceleration was associated with better memory test performance, which further supports the beneficial role of physical exercise on cognitive health.

Irisin browning in white adipose tissue increases uncoupling protein 1 levels in white fat cells, and circulating irisin in nanomolar levels enhance energy expenditure, improve glucose tolerance, and possess anti-obesity and anti-diabetic features (Bostrom et al., 2012). Irisin ablation in mice caused poor browning, hyperlipidemia, insulin resistance, reduced HDL levels, and poor bone strength (Luo et al., 2020). We found age-related decreases in irisin levels, which was attenuated by exercise training. We found circulating blood irisin is closely related to HDL concentration which has previously been reported (Rana et al., 2014; Sanchis-Gomar et al., 2014; Luo et al., 2020), but the link between irisin to GrimAge Acceleration and FitAge Acceleration is a novel observation. This work further supports the biological importance of irisin to the aging process and aligns with the relationship found between irisin and telomere length (Rana et al., 2014). It is possible our research motivates interventions to boost irisin, like through physical exercise, as possible anti-aging therapies.

The microbiomal pathways, which are associated with VO2max, could lead to increased availability of nitric oxide (NO), through the oxidation of L-arginine. Indeed, it has been shown that L-arginine would reduce O2 cost of running and improve endurance capacity (Jones et al., 2018) and this fits very well to the correlation between arginine biosynthesis promoting bacterial strains and VO2max. However, this could be the first report that observed the relationship between microbiotal L-arginine biosynthesis pathway and VO2max. Redox regulation is involved in almost all cellular processes, and redox regulation associated aging has mounting evidence (Radak et al., 2008b). L-rhamnose biosynthesis is correlated with redox balance potentially because l-rhamnose is an important component of glycan lipopolysaccharides of gram-negative bacteria. Therefore, it cannot be excluded that the microbiome is involved in the regulation of inflammatory and redox pathway through gut bacteria-derived proinflammatory nucleotide sugars. VO2max is negatively associated with microbiome pathways responsible for ornithine degradation (p= 0,000285057). This is just in accordance with the result of study which showed that L-ornithine supplementation decreased the appearance of fatigue during cycling due to promotion lipid metabolism and activated the urea cycle from serum triacylglycerol, ketone bodies, free fatty acids, and blood ammonia level changing (Sugino et al., 2008).

It was found that fecal microbiome pathways are heavily involved in the control of redox balance by pyruvate fermentation, and menaquinol.11.biosynthesis. When pyruvate is fermented, it is converted to lactate by lactate dehydrogenase, while NADH is oxidized to NAD+. NAD+/NADH ratio is used as redox marker of the cells, and our data suggest that microbiome pathways are taking part in the redox regulation state of humans. Menaquinone is an essential vitamin as an obligatory component of the electron transfer pathway in microorganisms and it appears that menaquinone might serve as an additional redox cofactor to mediate the proton-coupled electron transport across the membrane (Guan et al., 2018). HIGH-FIT microbiome bacterial flora support glycolysis, inosine biosynthesis, NAD salvage, and acetyl-CoA fermentation pathways to a greater extent than MED-LOW-FIT individuals.

PhenoAge acceleration was negatively associated with the degradation of D. galacturonate, D.fructuronate, and D.glucuronosides, and the end product of these pathways is pyruvate. This could suggest that lower levels of pyruvate are associated with PhenoAge Acceleration. In other words, more pyruvate may help decelerate DNAmPhenoAge. Pyruvate has a number of health promoting properties. It serves as energy-yielding metabolic fuel, it is a powerful anti-inflammatory agent, and it is an effective scavenger of hydrogen peroxide (Mallet et al., 2005; Das, 2006). PhenoAge has a strong relationship with the bacterial pathway of glyoxylate cycle, which promotes the production of glucose from fatty acids (Lorenz and Fink, 2002) providing fuel to the community of various bacterial species in the microbiome, emphasizing the importance of healthy microbiome in aging process.

DNAmFitAge is linked to mannosylglycerate biosynthesis pathway in the microbiome. The exact role of mannosylglycerate in mammals, however it is suggested that it is highly efficient in the protection of enzymes against thermal inactivation (Ramos et al., 1997). FitAge Acceleration was associated with isoleucine synthesis from threonine. Isoleucine is a branched-chain amino acid (BCAA) used for the biosynthesis of proteins. The level of BCAA is correlated with the survival of patients suffering from sepsis (Reisinger et al., 2021). Moreover, it has been shown that isoleucine level decreases with aging in the blood (Chaleckis et al., 2016), suggesting it is important to continually synthesize this amino acid to potentially mitigate aging phenotypes. Our research provides the first investigation between microbiome derived metabolic pathways and DNAm based aging biomarkers.

Through this complex investigation, we aim to gain a deeper understanding of how physical fitness is related DNAm based aging. We examined the relationship of physical fitness in healthy adults and life-long athletes to different physiological measurements, biochemical tests, memory tests, and DNAm based biomarkers. The newly created DNAmFitAge outperforms the existing DNA methylation based biomarkers and shows regular exercise is associated with younger biological age, better memory, and more protective blood serum levels. Based on these relationships DNAmFitAge could be an important biological marker of the quality of life.

## Data Availability

All data produced in the present study are available upon reasonable request to the authors

https://tf.hu/en/about-us/organization/departments/research-center-for-molecular-exercise-science

## Founding

ZR, FT, and MJ acknowledge support from the National Excellence Program (126823) and the Scientific Excellence Program, TKP2020-NKA-17, TKP2021-EGA-37 at the Hungarian University Sport Science, Innovation and Technology Ministry, Hungary. SH, KMM, and ATL acknowledge support from 1U01AG060908. KJAD was supported by grant # ES003598 from the National Institute of Environmental Health Sciences of the US National Institutes of Health, and by grant # AG052374 from the National Institute on Aging of the US National Institutes of Health.

